# Effects of latency on estimates of the COVID-19 replication number

**DOI:** 10.1101/2020.04.07.20056960

**Authors:** Lorenzo Sadun

**Affiliations:** Department of Mathematics, University of Texas, 2515 Speedway, RLM 8.100, Austin, TX, USA

## Abstract

It is not currently known how long it takes a person infected by the COVID-19 virus to become infectious. Models of the spread of COVID-19 use very different lengths for this latency period, leading to very different estimates of the replication number *R*, even when models work from the same underlying data sets. In this paper we quantify how much varying the length of the latency period affects estimates of *R*, and thus the fraction of the population that is predicted to be infected in the first wave of the pandemic. This variation underscores the uncertainty in our understanding of *R* and raises the possibility that *R* may be considerably greater than has been assumed by those shaping public policy.

---

A key step in understanding the spread of the COVID-19 pandemic is estimating the reproduction number *R*, namely the average number of new individuals that each infected individual goes on to infect. However, published estimates of the *basic* reproduction number *R*_0_, representing the initial value of *R* before actions were taken to reduce spread, have varied tremendously, from 1.3 (*1*) to more than 6 (*2*), and with non-overlapping error bars. The most widely cited estimate of *R*_0_ = 2.2 is that of Li et al (*3*), and most modeling of the future course of the pandemic has used Li’s value or lower.

Initial estimates of *R*_0_ were based on the early stages of the epidemic in Wuhan, China, where Li et al estimated a doubling time of 7.4 days. In situations where efforts have already been made to contain the spread, the actual reproduction number *R* should be less than *R*_0_; the goal of social distancing is to bring *R* below 1. However, despite public health measures, outbreaks in many places have grown much faster than in Wuhan. For instance, in the United States between March 8 and March 24, 2020, the number of confirmed COVID-19 cases grew 100-fold, from 541 to 54,856, corresponding to a doubling time of only 2.4 days (*7, 8*). Over the same time period, the number of confirmed cases in the entire world, excluding China, grew from 29,256 to 341,356, with a doubling time of 4.5 days; slower than the United States but faster than Wuhan.^1^

This paper addresses another point of confusion. While there is general agreement that some infected individuals who are not yet symptomatic (or who never develop symptoms) can still infect others (*4*), there is no agreement on how long it takes an infected individual to become infectious. Some researchers treat infected individuals as immediately infectious (*1, 5*), while others assume that they only become infectious towards the end of the incubation period (*6*). These different assumptions lead to different trajectories for a given *R*_0_, with earlier infectivity usually leading to faster growth. Conversely, when estimating *R*_0_ from an actual observed trajectory, models with earlier infectivity usually give lower estimates than models with a longer latency period. In this paper we quantify how very different those estimates can be.

To be reliable, estimates of *R* must be made for specific places and times and must acknowledge the large effects of the sometimes-arbitrary choices involved in model-building. We know much less than we think we know, and there is a substantial chance that the true value of *R* is much higher than the optimistic 1.3–2.5 range that has been assumed for most public policy discussions.

## Stage models

Most models of disease spread are variants of the SEIR model, which is itself an extension of the popular SIR model (*9*). In the SEIR model, people are classified as “Susceptible”, “Exposed”, “Infectious”, and “Removed” (or “Recovered”). Susceptible individuals become Exposed from contact with existing Infectious individuals, Exposed individuals become Infectious after a latency period, and Infectious individuals eventually recover, die, are quarantined, or are otherwise Removed from circulation. We let *S*(*t*), *E*(*t*), *I*(*t*) and *R*(*t*) denote the number of Susceptible, Exposed, Infectious and Removed individuals at time *t* and write equations to govern the evolution of these quantities.

It is important to acknowledge that even the SEIR model is not a complete description of disease spread. Now that the whole world is on alert, most symptomatic individuals are removed from circulation quickly. However, some show mild symptoms and are not noticed or removed from circulation, while some symptomatic individuals defy isolation orders or require a degree of home care that makes true isolation impossible. More accurate predictions require incorporating all of these phenomena (and others) correctly, which requires more complicated models, such as (*5*).

However, the limitations of the SEIR model are not the point of this paper. Our purpose here is to show that, even in the simplest models, results are highly dependent on the length of the Exposed state. This problem is not fixed by adding additional details to the model. On the contrary, the more complicated a model is, the more its results depend on the choices made in constructing that model.

The basic SEIR model has three continuous parameters.

1. The mean time *t*_1_ spent by patients in the uninfectious “exposed” state. We call this time the latency.
2. The mean time *t*_2_ spent by patients in the infectious state.
3. The reproduction number *R*, which is the mean number of people that each infected person infects in turn.

The sum *t*_*tot*_ = *t*_1_ + *t*_2_ is the total average time from exposure to the appearance of symptoms severe enough to cause an individual to isolate. We call this the extended incubation period. *t*_*tot*_ is relatively easy to measure, and for COVID-19 is believed to be about one week (*10*). The individual numbers *t*_1_ and *t*_2_ are much harder to determine. In this paper we will compare different values of the latency *t*_1_, while holding the sum *t*_*tot*_ fixed.

In addition to these three parameters, we consider several ways to treat the distribution of incubation times for individuals. The most common choice is to assume that the time each individual spends in the exposed state and the infectious state are independent exponential random variables with means *t*_1_ and *t*_2_, respectively. This choice leads to a simple system of differential equations. However, in this approach some exposed individuals move to the infectious stage almost immediately, which is not realistic. Alternatively, we can assume that the time spent in the exposed state is always exactly *t*_1_, and the time spent in the infectious state is exactly *t*_2_, but that ignores the fact that some people get sick much faster than others, while some people stay undetected and infectious for much longer than others. A promising approach is a hybrid, keeping the time in the exposed state constant and making the time in the infectious state random.

In the early stages of an outbreak, when almost all individuals are susceptible, these models (and more complicated extensions of these models) are all linear. Linearity implies that the total number of infected individuals grows exponentially as *e*^*t*/*τ*^, where *τ* is the doubling time of the growth divided by ln(2) ≈ 0.69. Since all models lead to the same qualitative behavior, *τ* is the only quantity that can be *directly* deduced from the data. The subsequent step of inferring *R* from *τ* then depends on which model we choose and which parameters we choose for that model.

## Results

We will concentrate on understanding the rapid growth of COVID-19 in the United States in March 2020, when *τ* was approximately 3.5 days. We apply each of the three models (exponential distribution of latency and the time spent infective, fixed times, and the hybrid) to compute *R* assuming that *τ* = 3.5 days and that the average *t*_2_ of the time spent infective is 7 days minus the average *t*_1_ of the latency.

The results are shown in Figure 1, which gives the computed value of *R*, for all three models, as a function of the latency *t*_1_. The relevant formulas are as follows: When both incubation times are treated as exponential random variables,

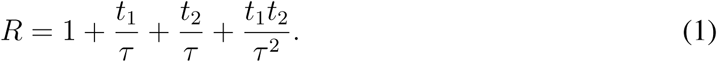

When both incubation times are fixed,

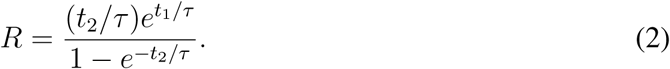

Finally, in the hybrid model

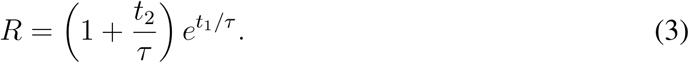

These formulas are derived in the Supplementary Materials.

**Figure 1:**
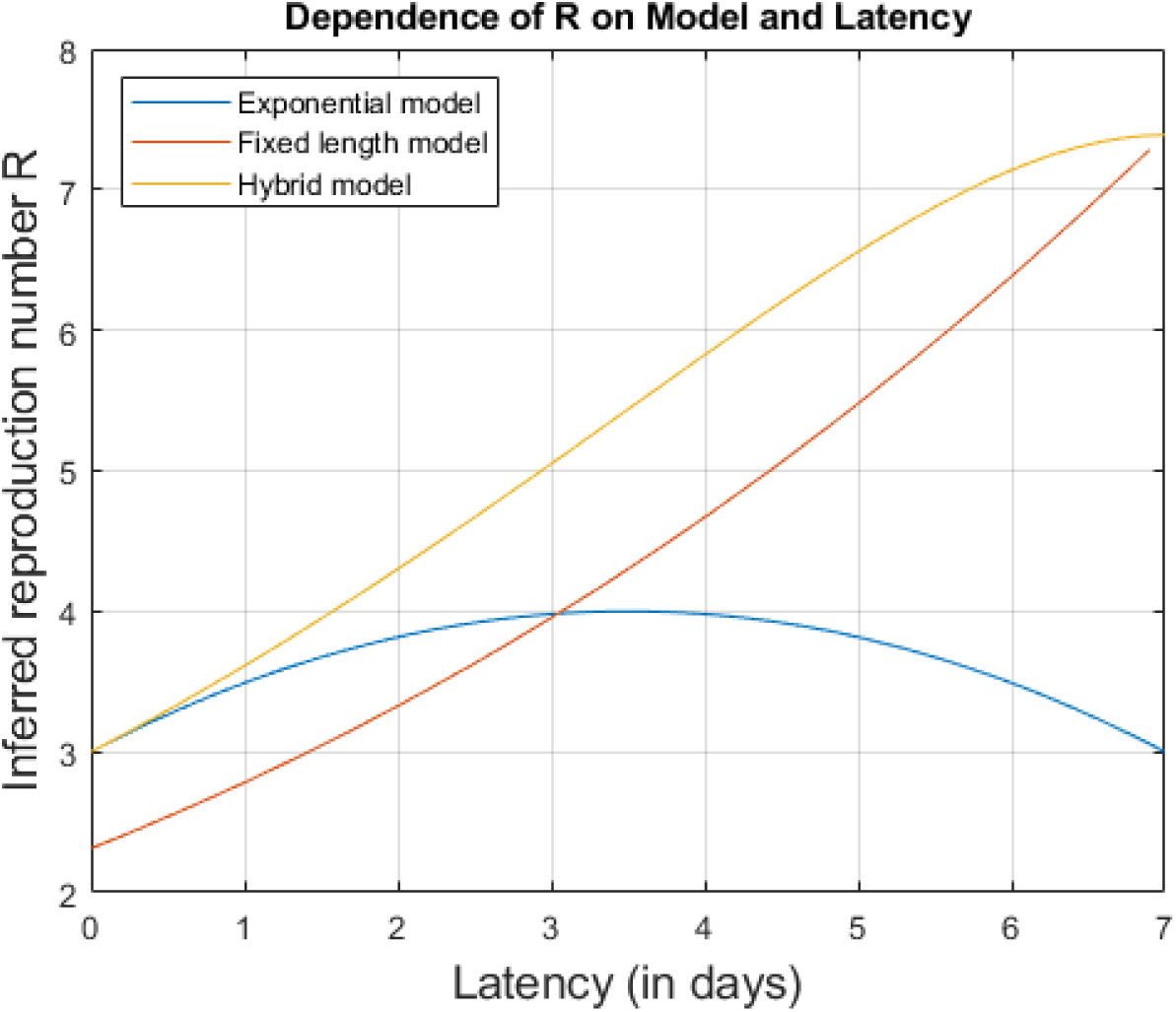
Estimated values of *R* for all three models as a function of the average latency *t*_1_. While increased latency greatly increases *R* in the fixed length and hybrid models, the exponential model behaves differently. The exponential model allows a few exposed individuals to reinfect others very quickly and these rapid transmitters drive much of the growth of the outbreak.

It is worth noting, not only how the inferred value of *R* depends on the average latency *t*_1_, but also how it depends on the seemingly innocuous choice of whether to make the lengths of the Exposed and Infectious stages fixed or random. While none of the models fit what is known about virology exactly, it is easy to make a good case for each of them. Yet they give vastly different results, especially for larger values of *t*_1_! Without a clear scientific justification for choosing one model over the other two, we must acknowledge that all of our estimates of *R* are extremely uncertain.

The exponential model behaves differently from the other models because it involves some individuals becoming infectious very quickly, even when the average latency is large, and involves some of those individuals infecting others very quickly as well. These rapid transmitters do not account for the majority of the transmissions, which still take a total of 7 days on average, but they have an oversized effect on the growth rate. The number of these rapid transmitters is maximized when either *t*_1_ is small or *t*_2_ is small, which explains why *R* reaches its peak when *t*_1_ = *t*_2_ = 3.5 days.

The lesson is that when the *minimum* time from infection to reinfection goes up, so does *R*. There is a simple explanation for this. If *τ* = 3.5, then the outbreak doubles every 2.4 days, quadruples every 4.8 days, and multiplies by 8 every 7.2 days. If it takes at least 4.8 days for an exposed individual to infect somebody else, then *R* must be at least 4 to account for the quadrupling in that time; if the time interval is always at least 7.2 days, then *R* must be at least 8.

The value of *R* has profound effects on public policy decisions. Suppose that a social distancing policy can reduce transmission by a factor of 3. If we start at *R* = 2, dividing *R* by 3 is enough to contain and eventually suppress the outbreak. If we start at *R* = 4, dividing by 3 will only slow the growth. In that case, more extreme measures are needed to get the outbreak under control.

*R* also determines the fraction of the population that eventually becomes exposed to the virus. In all of these models, that fraction *x* is the non-zero solution to the equation

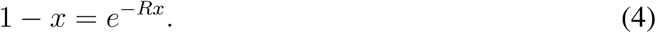

The dependence of *x* on *R* is shown in Figure 2. When *R* is as small as 1.15, *x* is already at 25%, and when *R* = 1.7, *x* is already about 70%. When studies such as (*11*) say that 25%– 70% of the population would be infected in the absence of control measures, they are implicitly assuming that *R*_0_ is only between 1.15 and 1.7. Under that assumption, moderate efforts should be enough to bring *R* below 1 and suppress the outbreak. However, if *R*_0_ is actually larger, and if our social distancing efforts don’t bring *R* below 2, then the vast majority of the population would be infected in the first wave of the pandemic. The resulting herd immunity would be impressive, but almost no vulnerable people would be left to benefit from it.

**Figure 2:**
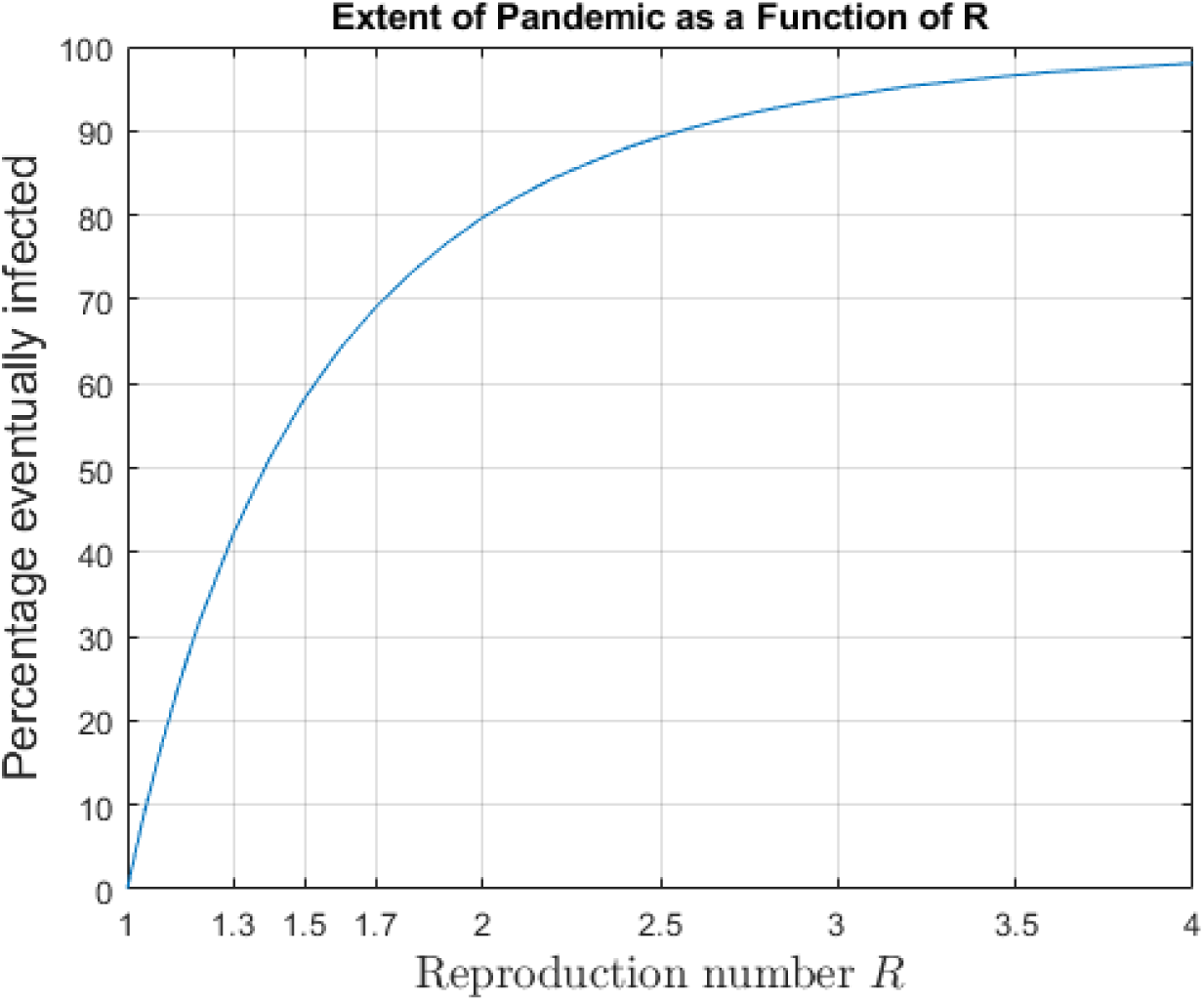
The percentage of the population that eventually becomes infected as a function of the reproduction number *R*. Note that this percentage is already over 40% when *R* = 1.3, and rapidly approaches 100% as *R* increases.

Finally, the value of *R* affects the possibility of having multiple waves of the pandemic. Suppose that a fraction *x* of the population is infected in the first wave, after which life returns to normal and *R* returns to its pre-lockdown value. Unless

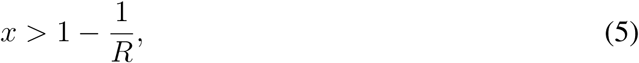

successive outbreaks are extremely likely to occur.

## Conclusions

All models for understanding the future trajectory of the COVID-19 pandemic depend on knowing how contagious the disease is; that is, on estimating *R*. This is true both for complicated models involving many different sorts of people and many different stages of disease progression, as well as for simple models such as SEIR. Unfortunately, there is no direct way to measure *R*. All we can do is measure the time scale *τ* of the exponential growth of the pandemic and try to estimate *R* from *τ*.

Such estimates depend strongly on the details of the model used. Eventually, the correct parameters and the most accurate models will be revealed through clinical studies of the actual distribution of the different stages of infection and especially by tracing individual contacts to determine when infectivity starts. Until those studies are completed for COVID-19, every projection will involve a choosing parameters through educated guesswork.

Even in the simple models considered in this paper, changing the parameters of the model can change the estimated value of *R* dramatically. In the fixed length model the estimate for *R* ranges from 2.3 to 7.4 and the hybrid model is almost as sensitive. In all three models, having a latency of just one day (*t*_1_ = 1) increases *R* by 0.5 or more compared to having no latency at all (*t*_1_ = 0). Working with more realistic (and more complicated) models only makes the problem of model dependence worse. The more parameters there are, the more the final answers depend on the assumptions.

As of early April 2020, most of the highly cited estimates of *R* have been based either on models without latency, on data from Wuhan that shows much slower growth than seen in Europe and the United States, or both. As such, these estimates are likely to have substantially underestimated the true value of *R* in the United States.

The success of any suppression strategy boils down to one thing: *getting R below 1*. Strategies that could work if *R* is between 1.3 and 2.5 might well fail if the actual value of *R* is larger. Meanwhile, mitigation strategies depend on isolating the most vulnerable members of society until the first wave of the pandemic has subsided. The amount of herd immunity required to make that work also depends on *R*. It won’t be safe for the vulnerable to go out until the first wave of the pandemic has subsided *and* a fraction 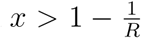 of the population has developed immunity through exposure or vaccine. Higher values of *R* provide a stronger argument for accelerating the testing of a vaccine and accepting the accompanying risks of adverse side effects (by rushing Phase I) or settling on a less effective vaccine (by rushing Phase II). Haste may well be needed.

The world has put enormous effort and expense into confronting COVID-19. To succeed in the long term, we need a much better understanding of the infectivity of COVID-19. Models with accurate descriptions of what happens after incubation can help, but only after we have a much better understanding of the minimum time between infection and infectivity. Without that understanding, all of our models will remain unreliable and our policies will remain uninformed.

## Data Availability

All data is obtained from public sources listed in the references.

## Acknowledgements

I thank Tony Moody, Stan Schein, Alfredo Sadun, Anita Sadun, Rebecca Sadun and Rina Sadun for their enormous help in making this paper comprehensible to a non-mathematical audience. This work is supported by the University of Texas at Austin, but not through any external grants. There are no conflicts of interest. All data is available in the manuscript or the supplementary materials.

## Supplemental materials: Mathematical derivations

All versions of the SEIR model have the same general structure.

1. Susceptibles turn into Exposed at a rate equal to *βS*(*t*)*I*(*t*), where *β* is a transmission coefficient. In the early stages of the pandemic, *S*(*t*) is approximately the entire population *T*, so the number of Susceptibles who become Exposed, per unit time, is approximately *βTI*(*t*).
2. Exposed individuals become Infectious in an average of *t*_1_ days. In the exponential version, the number of Exposed who become Infectious per unit time is *αE*(*t*), where *α* = 1/*t*_1_. In the fixed-length version, the rate at which Exposed become Infectious today is exactly equal to the rate at which they became Exposed *t*_1_ days ago, namely *βTI*(*t*−*t*_1_).
3. Infectious individuals are Removed in an average of *t*_2_ days. In the exponential version, the number of Infectious who are Removed per unit time is *γI*(*t*), where *γ* = 1/*t*_2_. In the fixed-length version, the rate at which Infectious are Removed today is exactly equal to the rate at which they became Infectious *t*_2_ days ago, which in turn is the rate at which they were Exposed *t*_1_ + *t*_2_ days ago, namely *βTI*(*t* − *t*_1_ − *t*_2_).
4. The reproduction ratio is *R* = *t*_2_*βT*, so we can replace *βT* with *R/t*_2_ or *Rγ*.

### Exponential incubation times

In the purely exponential model, these considerations yield the following system of differential equations:

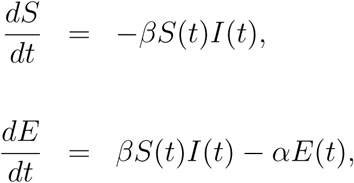

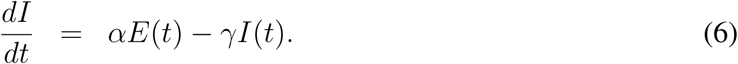

Making the approximation *βS*(*t*) ≈ *βT* = *Rγ*, we get

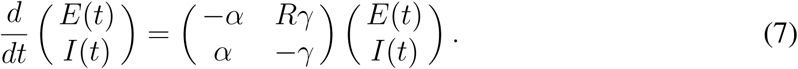

The exponential rate of growth (1/*τ*) is the positive eigenvalue of the matrix on the right hand side. Since the sum of the two eigenvalues is the trace, namely −(*α* + *γ*), the other eigenvalue must be 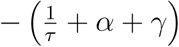. The determinant of the matrix is −(*R* − 1)*αγ*. Since the determinant is the product of the two eigenvalues, we have

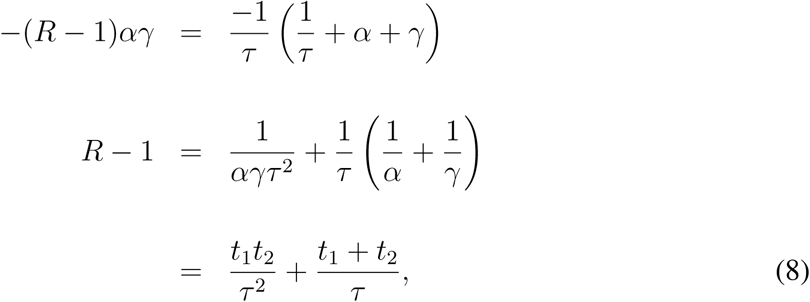

which is equivalent to equation (1).

### Fixed incubation times

In the model with fixed incubation times, Exposed become Infected at rate *RI*(*t* − *t*_1_)/*t*_2_, while Infected are Removed at rate *RI*(*t* − *t*_1_ − *t*_2_)/*t*_2_, so

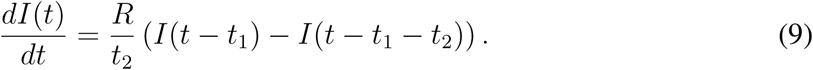

Plugging in *I*(*t*) = *Ce*^*t/τ*^, where *C* is an unknown constant, we obtain

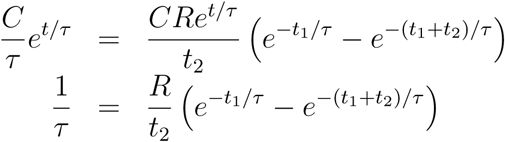

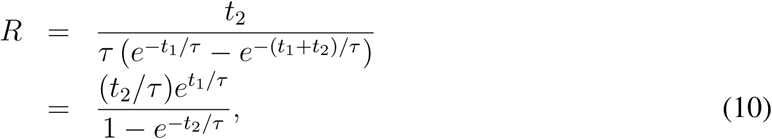

which is equation (2).

### The hybrid model

Here we treat the length of the exposed stage as fixed and the length of the infectious stage as random. The rate at which Exposed become Infectious is *RγI*(*t* − *t*_1_) = *RI*(*t* − *t*_1_)/*t*_2_, but the rate at which Infectious individuals are removed is *γI*(*t*) = *I*(*t*)/*t*_2_. This gives the differential equation

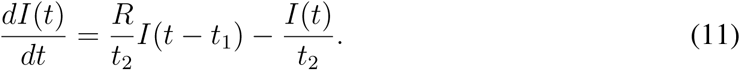

Plugging in *I*(*t*) = *Ce*^*t/τ*^ as before, and dividing both sides by *Ce*^*t/τ*^, gives

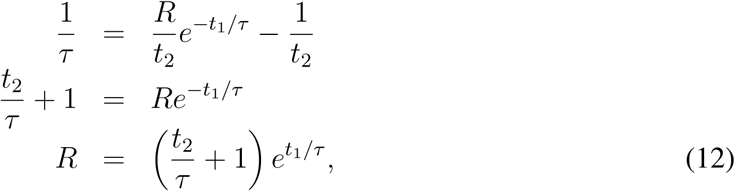

which is equation (3).

### How many people will eventually get sick?

Finally, we consider what fraction of people will get sick before the pandemic ends. The pandemic burns itself out when it runs out of Susceptibles to infect, so for this part of the calculation we do *not* make the simplifying approximation that *S*(*t*) ≈ *T*. Instead we look at the exact equation for *S*, which is the same in all versions of our model.

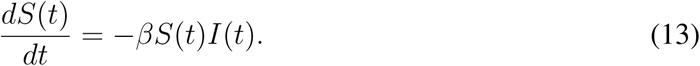

Dividing both sides by *S*(*t*) gives

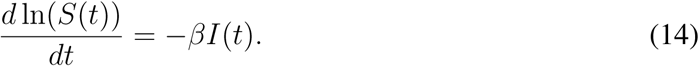

Integrating then gives

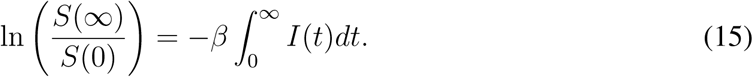

Let *x* be the fraction of people who eventually get sick. The left hand side of equation (15) is ln(1 − *x*), while the right hand side is −*βt*_2_*S*(0)*x*, since the number of people who get sick is *xS*(0), and since each sick person is infectious for an average of *t*_2_ days. However,

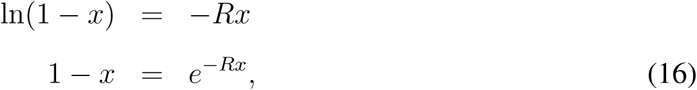

which is equation (4).

### Multiple waves and herd immunity

To understand equation (5), imagine that *R* = 4, so each infected person has contact, of the sort that spreads the virus, with an average of 4 other people. If more than 3/4 of the population has already had the virus, then fewer than one of those four people (on average) will get infected. Since each infected individual actually infects fewer than one new person (on average), any local outbreak will quickly peter out.

More generally, if a fraction *x* of the population is already immune and a fraction 1 − *x* is still Susceptible, the average number of new people that each infected individual actually infects is only *R*(1 − *x*), not *R*. As long as *R*(1 − *x*) < 1, or equivalently 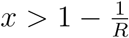, local outbreaks cannot get any traction and cannot spread into the population at large. This phenomenon, where immunity in part of the population protects the rest, is called “herd immunity”.

### Footnotes

1 These numbers are for confirmed cases. Due to testing inefficiencies, the actual number of cases on March 8 was probably much higher than reported, and likewise on March 24. However, if the number of cases in the United States only grew 50-fold instead of 100-fold, the resulting doubling time of 2.8 days would still be far smaller than in Wuhan.

